# GLUCOSE: A Distributional Reinforcement Learning Model for Optimal Glucose Control After Cardiac Surgery

**DOI:** 10.1101/2025.01.01.25319851

**Authors:** Jacob M. Desman, Zhang-Wei Hong, Moein Sabounchi, Ashwin S. Sawant, Jaskirat Gill, Ana C Costa, Gagan Kumar, Rajeev Sharma, Arpeta Gupta, Paul McCarthy, Veena Nandwani, Doug Powell, Alexandra Carideo, Donnie Goodwin, Sanam Ahmed, Umesh Gidwani, Matthew Levin, Robin Varghese, Farzan Filsoufi, Robert Freeman, Avniel Shetreat-Klein, Alexander W Charney, Ira Hofer, Lili Chan, David Reich, Patricia Kovatch, Roopa Kohli-Seth, Monica Kraft, Pulkit Agrawal, John A. Kellum, Girish N. Nadkarni, Ankit Sakhuja

## Abstract

This study introduces Glucose Level Understanding and Control Optimized for Safety and Efficacy (GLUCOSE), a distributional offline reinforcement learning algorithm for optimizing insulin dosing after cardiac surgery. Trained on 5,228 patients, tested on 920, and externally validated on 649, GLUCOSE achieved a mean estimated reward of 0.0 [-0.07, 0.06] in internal testing and -0.63 [-0.74, -0.52] in external validation, outperforming clinician returns of -1.29 [-1.37, -1.20] and -1.02 [-1.16, -0.89]. In multi-phase human validation, GLUCOSE first showed a significantly lower mean absolute error (MAE) in insulin dosing, with 0.9 units MAE versus clinicians’ 1.97 units (p < 0.001) in internal testing and 1.90 versus 2.24 units (p = 0.003) in external validation. The second and third phases found GLUCOSE‘s performance as comparable to or exceeding that of senior clinicians in MAE, safety, effectiveness, and acceptability. These findings suggest GLUCOSE as a robust tool for improving postoperative glucose management.

## INTRODUCTION

Cardiac surgery elicits a substantial metabolic stress response resulting in postoperative hyperglycemia regardless of diabetic status (1). Post-operative hyperglycemia after cardiac surgery is common, occurring in 60-80% patients with diabetes (2), and over 50% non-diabetic patients (3). It is associated with higher rates of post-operative infections (4, 5), acute kidney injury (3, 6–8), cardiac arrhythmias (3), longer length of stay (3), and higher mortality (6–9). Due to its significance, the Society of Thoracic Surgeons (STS) recommends maintaining blood glucose levels below 180mg/dL after cardiac surgery (10).

Achieving adequate glucose control post-operatively is challenging. A study found that only 15% of patients had appropriate glucose control, defined as glucose level between 70mg/dL to 180mg/dL, within the first day after cardiac surgery (11). This early post-operative period, when patients are critically ill and require care in intensive care unit (ICU) settings, is highly dynamic with rapidly changing clinical characteristics of patients. Currently, post-operative glucose management involves titration of regular insulin based on hospital specific protocols and the experience of treating clinicians.

However, due to the highly dynamic nature of this early post-operative period, some treatment regimens may be more suitable for certain patients or only effective for a limited time as their condition evolves. This leads to high rates of hyperglycemic and hypoglycemic episodes (11, 12), both associated with worse outcomes, as these protocolized regimens often fail to account for individual patient variability in real-world settings (13, 14). Therefore, personalized and dynamic insulin titration is crucial for improving glucose control in patients following cardiac surgery.

Reinforcement Learning (RL), a type of machine learning where an agent learns to make decisions by performing actions in an environment to maximize cumulative rewards, offers a promising solution to this challenge (15). RL algorithms receive feedback in the form of rewards or penalties based on the actions taken, allowing the agent to improve its policy over time. This adaptability makes RL particularly well-suited for tasks that involve complex decision-making and require real-time adjustments, such as insulin titration in the dynamic postoperative environment.

Implementing an RL-based system for insulin titration can address the limitations of current glucose management protocols. By continuously learning from individual patient data, RL can provide personalized treatment plans that account for specific patient variability and maintain glucose in optimal range. Additionally, RL’s capability to adapt to rapidly changing clinical characteristics ensures that insulin dosing remains optimal as patient conditions evolve. Traditionally, offline RL, where the agent learns from a fixed dataset without further interaction with the environment, has been limited by its focus on expected rewards, often overlooking the uncertainty in patient responses (16). This limitation can lead to suboptimal treatment plans, as it fails to account for the full spectrum of possible outcomes. As a result, offline RL systems may not adequately address the diverse risk profiles associated with different patient actions, potentially compromising the safety and effectiveness of interventions (17).

Our approach addresses this limitation by integrating distributional RL, which characterizes the entire distribution of potential outcomes rather than just the expected reward (17). This methodology provides a more comprehensive understanding of the risks and benefits associated with various actions, allowing for more nuanced decision-making under uncertainty (17–19). By considering the full range of potential patient responses, distributional RL can enhance the personalization and safety of insulin titration protocols, ensuring optimal dosing as patient conditions change.

Our proposed model, Glucose Level Understanding and Control Optimized for Safety and Efficacy (GLUCOSE), aims to improve glucose management on the first day after cardiac surgery, potentially leading to better patient outcomes and more effective clinical decision-making. We have developed GLUCOSE using data of patients undergoing cardiac surgery in the Medical Information Mart for Intensive Care-IV (MIMIC-IV) database (20). We then validated the model externally with cardiac surgery patients from the eICU Collaborative Research Database (eICU-CRD), a diverse, multicenter database of critically ill patients (21).

## RESULTS

### Study Population

GLUCOSE was trained and validated on two separate ICU datasets: the MIMIC-IV database (20) and the eICU-CRD database (21). MIMIC-IV was used as the development cohort and split into training and internal testing sets. eICU-CRD was used as the external validation dataset. Based on the inclusion and exclusion criteria, our study included 6,148 patients in development dataset and 649 patients in external validation dataset. The mean age of patients in the development dataset was 67.8 ± 11.6 years with 71.1% males, and in external validation dataset was 67.0 ± 11.3 years with 67.2% males. At least one hypoglycemic event (<70 mg/dL) occurred in 7.6% of patients in the development dataset and among 7.2% of patients in the external validation dataset. Similarly, at least one hyperglycemic event (>180 mg/dL) occurred in 47.8% of patients in development dataset and 47.3% of patients in external validation dataset. The baseline characteristics of the patients are shown in Table 1 and Supplementary Table 1. The overall structure of our study is illustrated in Fig. 1.

**Table 1.** Patient characteristics

|  |  | Overall | Development Cohort | External Validation Cohort | P-Value |
| --- | --- | --- | --- | --- | --- |
| Admission Age, median [Q1, Q3] |  | 68.0 [61.0,76.0] | 68.0 [61.0,76.0] | 67.0 [60.0,75.0] | 0.093 |
| Gender, n (%) | F | 1959 (28.8) | 1746 (28.4) | 213 (32.8) | 0.02 |
|  | M | 4838 (71.2) | 4402 (71.6) | 436 (67.2) |  |
| Height, median [Q1,Q3] |  | 173.0 [165.0,178.0] | 173.0 [165.0,178.0] | 170.2 [162.6,177.8] | 0.059 |
| Weight, median [Q1,Q3] |  | 83.6 [72.1,96.2] | 83.2 [72.0,96.0] | 85.6 [72.6,99.7] | 0.005 |
| BMI, median [Q1,Q3] |  | 28.3 [25.1,32.3] | 28.2 [25.1,32.1] | 29.2 [25.8,33.9] | <0.001 |
| 24h SOFA Score, median [Q1,Q3] |  | 5.0 [3.0,7.0] | 5.0 [3.0,7.0] | 6.0 [4.0,8.0] | <0.001 |
| Race, n (%) | ASIAN | 151 (2.2) | 136 (2.2) | 15 (2.3) | <0.001 |
|  | BLACK | 273 (4.0) | 222 (3.6) | 51 (7.9) |  |
|  | HISPANIC | 254 (3.7) | 175 (2.8) | 79 (12.2) |  |
|  | NATIVE | 11 (0.2) | 11 (0.2) |  |  |
|  | OTHER | 219 (3.2) | 219 (3.6) |  |  |
|  | UNKNOWN | 913 (13.4) | 841 (13.7) | 72 (11.1) |  |
|  | WHITE | 4976 (73.2) | 4544 (73.9) | 432 (66.6) |  |

**Figure 1.**
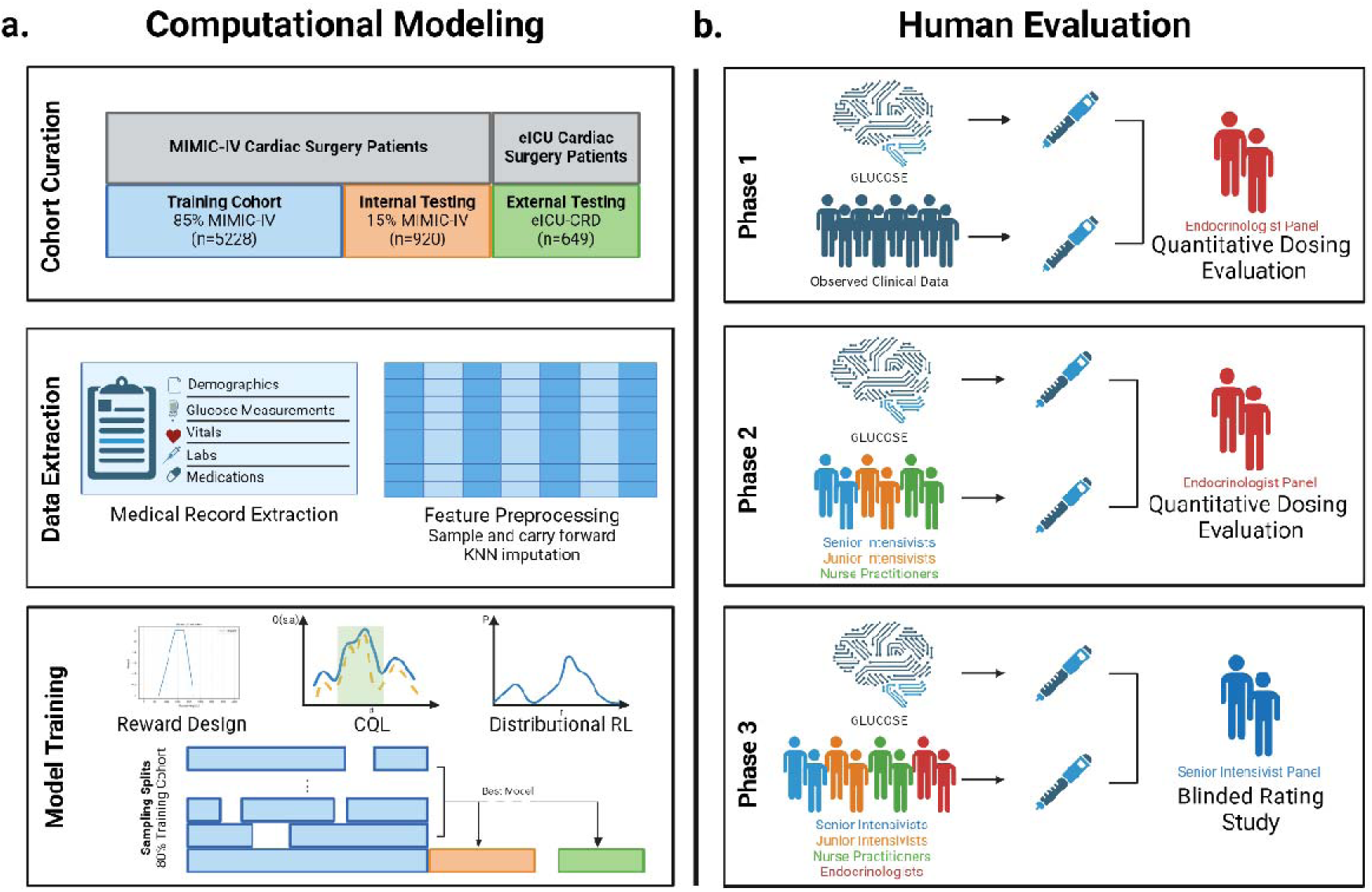
Study overview. **a)** Schema for model development, testing, and selection. **b)** Overview of clinician validation study.

### Performance of GLUCOSE

To mitigate the sampling and stochastic biases inherent in offline RL (22) we trained, in line with previous literature, multiple models until we observed no significant improvements in the RL policies (23, 24) (25). Consequently, we trained 200 independent models and selected the model with the maximal lower bound of the 95% CI of mean estimated performance returns within the internal testing set as the GLUCOSE model. We compared the estimated performance returns of GLUCOSE at the lower bound of its 95% CI with the upper bound of clinicians’ 95% CI (Fig. 1) using fitted Q estimation (FQE) for off policy evaluation (OPE) (26, 27), illustrating the differences in average estimated performance after evaluating 200 policies. The dotted blue and dotted orange lines reflect the 95% confidence intervals of the mean performance for the observed clinicians behavior in internal testing and external validation, respectively, while their non-dotted counterparts reflect the estimated performance of GLUCOSE through OPE (Fig. 2a). The best model, GLUCOSE, resulted in a mean estimated performance return of 0.0 [-0.07, 0.06] in the internal testing set and -0.63 [-0.74, -0.52] in the external validation dataset, showing significant improvements over the clinician returns of -1.29 [-1.37, -1.20] in the internal testing set and -1.02 [-1.16, -0.89] in the external validation dataset.

**Figure 2.**
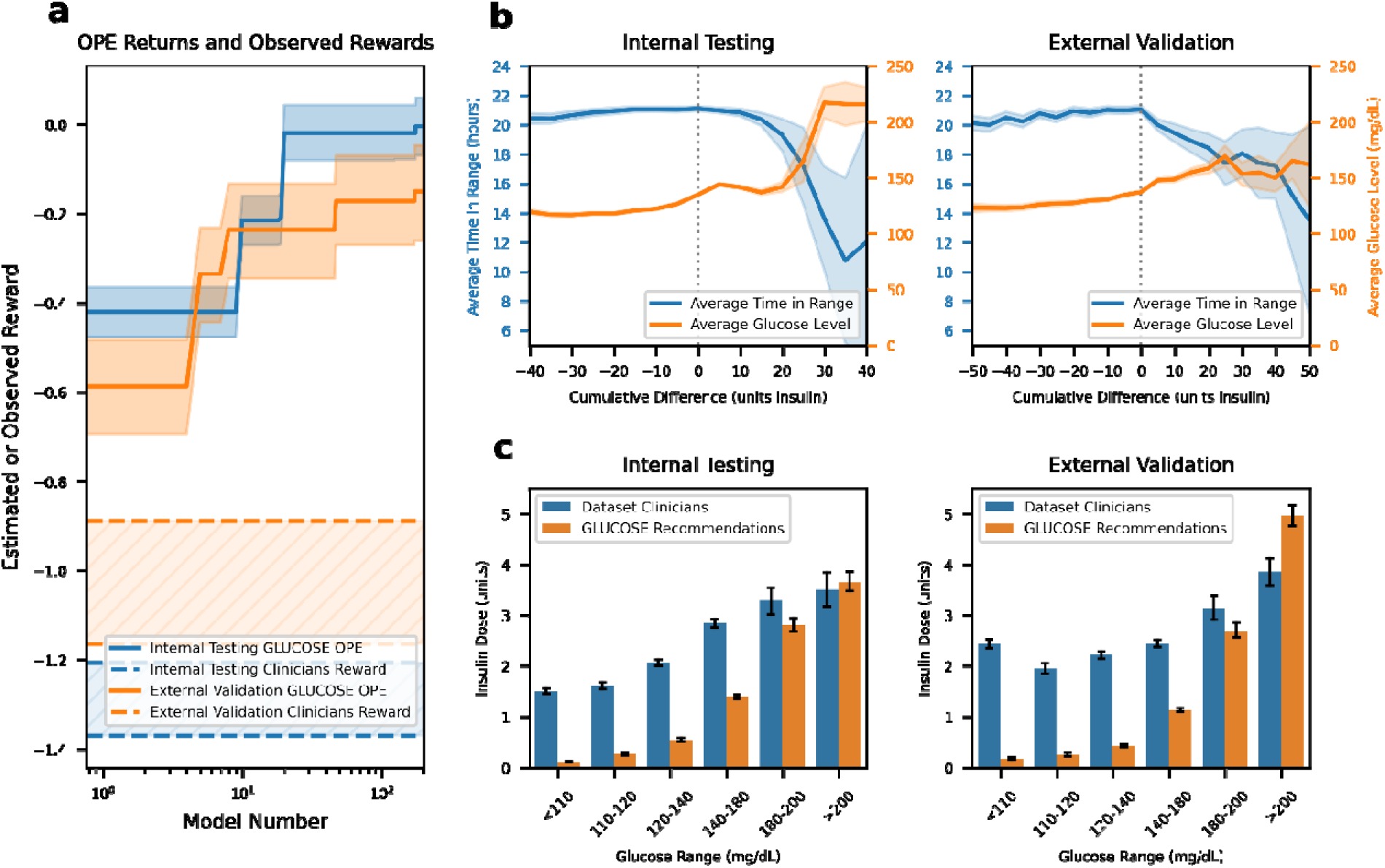
GLUCOSE performance. **a)** OPE counterfactual estimated performance of the model computed by FQE (solid lines) compared to the returns by the treating clinicians (dotted lines) with 95% CI. **b)** Comparison of TIR and average glucose relative to insulin dosing differences with 95% CI. **c)** Average insulin doses across several glucose ranges with 95% CI.

### GLUCOSE Policy Analysis

We further assessed the model by evaluating the time in range (TIR) of 70-180 mg/dL for glucose level when the actual clinician administered insulin dose was similar or different from the dose recommended by GLUCOSE. In the internal testing set, 27.3% of the time patients received insulin doses from clinicians identical to those recommended by GLUCOSE, while in the external validation dataset, this occurred 20.3% of the time. As shown in Fig. 2b, patients who received insulin doses like those suggested by GLUCOSE had the highest average TIR in both the internal testing set and external validation dataset. The TIR decreased as the difference of model recommended doses minus clinician administered doses increased, indicating that the model identifies areas for improvement in insulin management. For example, at more negative cumulative differences, where the average glucose is also lower, GLUCOSE suggests less insulin to mitigate the risk of hypoglycemia (Fig. 2b,c). Conversely, at more positive cumulative differences, where average blood glucose is higher and average TIR is worse, the model suggests higher insulin doses to avoid hyperglycemia. The action distribution of Fig. 2c further illustrates these trends, showing that insulin doses are lower on average at lower glucose levels and higher at higher glucose levels.

To gain insight into model representations and ensure its clinical interpretability, we derived feature importances for GLUCOSE using SHapley Additive exPlanations (SHAP) (Supplementary Fig. 1) (28). This analysis revealed that the most heavily weighted features align well with clinical intuition. The current glucose level emerged as the most critical feature, with recent glucose values from the previous 4 hours and weight also being among the highest contributors. Additionally, factors indicating patient acuity, such as the use and duration of mechanical ventilation, were weighted heavily. These findings suggest that GLUCOSE uses clinically relevant information in its decision making.

### Human Evaluations of GLUCOSE

For clinical applicability and robustness, we conducted a multi-phased human evaluation. In the first phase, two senior endocrinologists, each with over 10 years of clinical experience, provided their recommendations for hourly insulin dosing for the first day after cardiac surgery for 10 patients in both internal testing and external validation datasets. To allow the endocrinologists to provide the most accurate dosing schemes to use as a reference, we provided them with the entire time series of patient data, including the insulin doses actually administered by the treating clinicians, and resultant glucose levels. We compared the hourly insulin doses recommended by GLUCOSE, which unlike the endocrinologists had access only to the current state, to those actually administered by clinicians using the average endocrinologist doses as the reference. Across both datasets, GLUCOSE achieved significantly lower mean absolute error (MAE) in hourly insulin dosing, indicating that its dosing scheme more closely aligned with the recommendations of the endocrinologists. In the internal testing set, GLUCOSE had an MAE of 0.9 units compared to the treating clinician’s 1.97 units MAE (p < 0.001). In the external validation dataset, GLUCOSE had an MAE of 1.90 units compared to the treating clinician’s MAE of 2.24 units (p = 0.003).

In the second phase, two senior cardiac intensivists (>5 years’ experience), two junior cardiac intensivists (<5 years’ experience), and two cardiac intensive care unit nurse practitioners provided their recommendations for hourly insulin doses for the same patients. These clinicians were also provided with the entire time series of data, actual insulin administration record, and glucose levels to allow them to generate their most retrospectively optimal possible human policies. We then compared the GLUCOSE recommended doses, which again only had access to a single state of information at the current timestep, to those recommended by these 6 clinicians, with the endocrinologist recommendations as the reference (Fig. 3a). In internal testing, GLUCOSE achieved an MAE of 0.90 units compared to that of senior intensivists’ 0.82 unit MAE (p=0.57), junior intensivists’ 1.15 unit MAE (p=0.25), and nurse practitioners’ 1.23 unit MAE (p=0.21). In external validation, GLUCOSE achieved an MAE of 1.90 units compared to that of senior intensivists’ 1.58 units (p=0.32), junior intensivists 2.15 units (p=0.53), and nurse practitioners 2.28 units (p=0.38). Although the differences in MAE did not reach statistical significance, GLUCOSE demonstrates a trend toward lower MAEs than that of junior intensivists and nurse practitioners when compared against endocrinologists as the reference.

**Figure 3.**
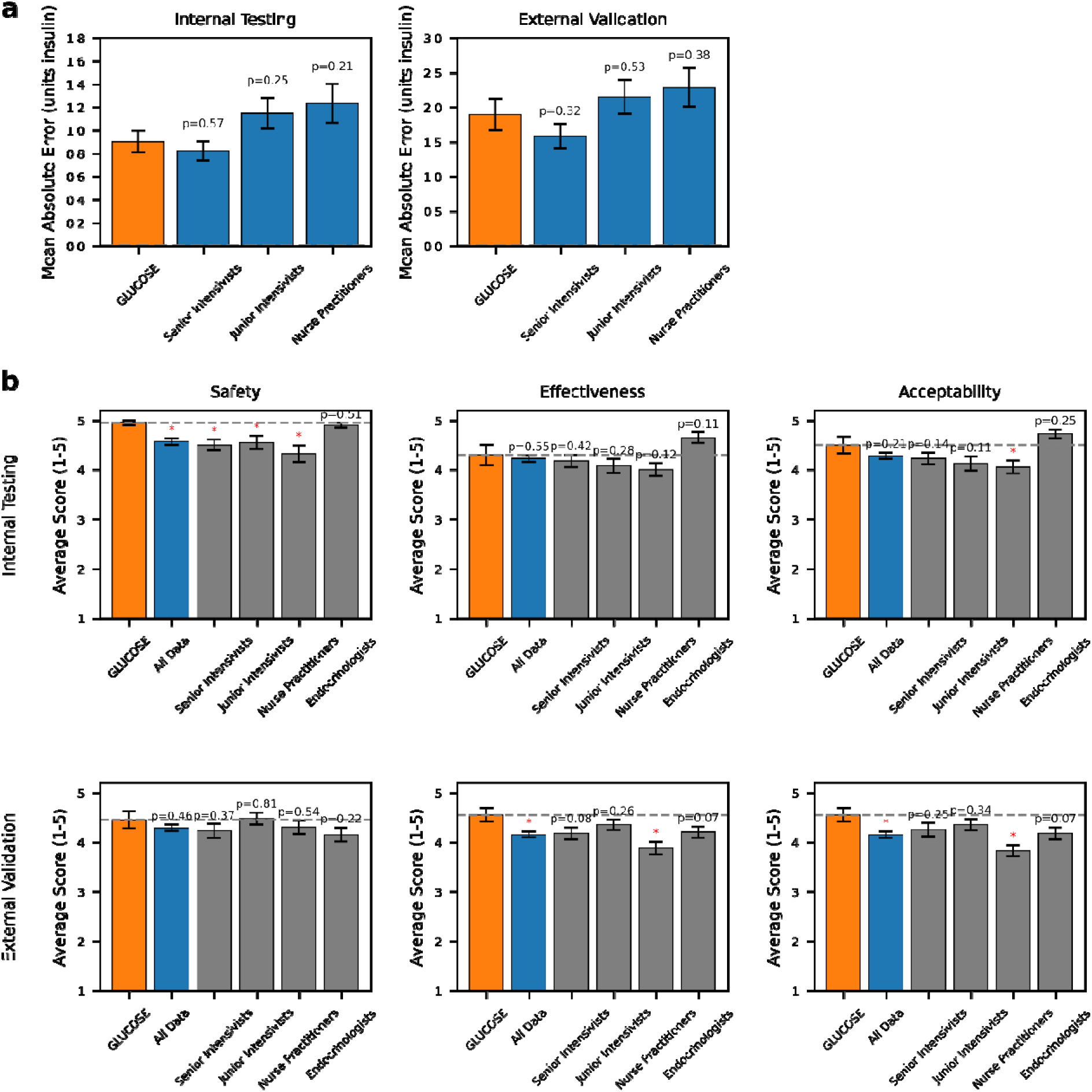
Human validation study results. **a)** MAE of clinician groupings and GLUCOSE relative to an endocrinologist baseline with standard error of the mean. **b)** Blinded ratings across safety, effectiveness, and acceptability of all clinicians by a blinded senior intensivist panel with standard error of the mean.

In the final phase, we conducted a blinded evaluation of GLUCOSE and all 8 clinician dosing recommendations using an expert panel of 2 separate senior intensivists to assess practical safety, effectiveness, and acceptability of the model’s recommended insulin doses. The two additional senior intensivists used a 5-point Likert scale to assess the safety (to reduce hypoglycemia), effectiveness (if the regimen would bring glucose into an acceptable range), and acceptability (if the regimen would be acceptable in a clinical scenario) of each recommended insulin regimen for the same group of 20 patients. In both internal testing and external validation datasets, GLUCOSE’s rated safety, effectiveness, and acceptability demonstrated either comparable performance or statistically significant improvements over all human policies (Fig. 3b). Notably, GLUCOSE performed at or above the level of senior cardiac surgery intensivists across all domains in both the internal testing and external validation datasets. This demonstrates GLUCOSE’s reliability and consistent high-level performance across diverse clinical scenarios.

### Evaluation of GLUCOSE’s Recommendations Among Excluded Patients

Finally, we evaluated GLUCOSE‘s recommendations in subsets of the external validation dataset that were excluded from the primary analysis due to the presence of ambiguous administration of insulin, vasopressors or inotropes. These patients had documented insulin, vasopressor, or inotrope administration but with insufficient information to determine the exact timing or dose - an issue commonly reported in the multicenter eICU-CRD database (29). Based on the affected medication, we performed this evaluation separately in patients with only ambiguous insulin data (3,001 patients) and in those with ambiguous data for both insulin and vasopressors/inotropes (1,804 patients). As there were only 33 patients with non-ambiguous insulin but ambiguous vasopressor/inotrope data, we excluded them from this analysis.

The external validation cohort, the ambiguous insulin subset, and the ambiguous medication subset were similar in terms of age, gender, and weight. However, these additional subsets included a higher proportion of white patients (66.6% vs 81.1% vs 89.7%, p < 0.001) and fewer patients with type 2 diabetes (9.1% vs. 3.1% vs. 1.2%, p < 0.001). While there were statistically significant differences in the average glucose levels (134.5 mg/dL vs. 132.2 mg/dL vs. 130.6 mg/dL, p < 0.001) these small differences are not clinically meaningful. Full demographic analysis can be found in Supplementary Table 2.

Due to the lack of accurately recorded insulin in these subgroups, we were unable to perform OPE or direct comparisons which depend on accurate insulin administration records. However, the overall distribution of model recommended actions was comparable across datasets (Supplementary Fig. 2). Although there were statistically significant differences, all differences in average insulin across all glucose ranges were less than half a unit and therefore not clinically significant (Supplementary Fig. 2).

## DISCUSSION

In this study we have developed GLUCOSE, a distributional offline RL based algorithm, that dynamically suggests personalized regular insulin dosing for patients in the first day after cardiac surgery. The algorithm was validated on an independent multicenter dataset and further demonstrated its robustness and safety through rigorous human evaluations.

Hyperglycemia early after cardiac surgery is associated with higher rates of post-operative infections (4, 5), acute kidney injury (3, 6–8), cardiac arrhythmias (3), longer length of stay (3), and higher mortality (6–9). This underscores the importance of glucose control in the early post-operative period. Moreover, research indicates that the harmful effects of hyperglycemia are dose-dependent, with longer exposure and higher glucose levels leading to worse outcomes (30). Therefore, it is essential to manage both the severity and duration of hyperglycemia. The STS recommends maintaining blood glucose levels below 180mg/dL after cardiac surgery (10). To achieve this target, most cardiac surgery centers employ institutional protocols for managing hyperglycemia (31–34). However, a significant challenge in early postoperative glucose management is that insulin, the primary treatment for hyperglycemia, has a narrow therapeutic window (35). Since these protocolized regimens often fail to account for individual patient variability in real-world settings (13, 36), hypoglycemia becomes a significant risk, particularly with intensive insulin dosing schemes (37, 38). Hypoglycemia, defined as a blood glucose level <70mg/dL, can trigger increased sympathetic activity leading to increased heart rate or arrhythmias (39), impairment of autonomic cardiac reflexes (40), poor neurological outcomes (41), and death (12). This hypoglycemia is seen in 5-21% patients after cardiac surgery (11, 12, 37) prompting a more conservative insulin dosing which, in turn, can result in persistent hyperglycemia. Thus, not surprisingly, these protocols frequently fall short, with only 15% of patients reaching the recommended glucose levels without hypoglycemia on the first day after surgery (11), which is the most critical and dynamic period after cardiac surgery.

Clinicians review over 1,300 data points per patient each day, making it difficult to effectively use all this information for clinical decision-making (42). An algorithm that can systematically process and interpret these data points can significantly enhance clinician workflow while improving patient outcomes. The GLUCOSE model addresses this by evaluating over 70 features, such as vasopressor doses, SOFA score, mechanical ventilation needs, past glucose values, and BMI, every hour. It recommends personalized insulin doses that account for the patient’s evolving clinical status. Importantly, the algorithm prioritizes features that are clinically relevant, as reflected in the feature importance analysis. Consistently, the GLUCOSE dosing scheme outperforms traditional clinician-driven dosing strategies in terms of estimated average performance.

The TIR for glucose was highest when the administered insulin dose closely matched the model’s recommendations. As the discrepancy between the administered doses and GLUCOSE’s suggested doses increased, the time in range decreased. Notably, when the difference was negative, meaning GLUCOSE recommended less insulin than what was administered, the average glucose level was lower. Conversely, with positive differences—where GLUCOSE suggested more insulin than what was given—the average glucose level was higher. This indicates that aligning insulin doses more closely with GLUCOSE’s recommendations could potentially increase time in range and reduce glucose variability, which is associated with worse clinical outcomes (13, 14).

Although there have been algorithms developed to assist clinicians in insulin doses, they are mostly limited to simulated settings without any human evaluations, include exclusively patients with diabetes, and none specifically target post-cardiac surgery patients (43–45). To ensure clinical applicability and acceptability of our study we performed a comprehensive 3-phase human evaluation inspired by prior work (43), which is a significant strength of our study. The results of our multi-phased human evaluation underscore the clinical robustness and reliability of the GLUCOSE algorithm in guiding insulin dosing for post-cardiac surgery patients. The significant reduction in MAE achieved by GLUCOSE compared to observed clinician dosing across both internal and external datasets highlights the algorithm’s agreement with rigorous clinical evaluation. Particularly noteworthy is GLUCOSE’s performance in the final phase of the evaluation, where it was assessed by senior intensivists on safety, effectiveness, and acceptability. The algorithm was either comparable to or exceeded the standards set by experienced clinicians, including senior cardiac surgery intensivists. It is important to note, that unlike GLUCOSE, which only had access to patient data till each current time-step, the clinicians that performed human evaluations had access to the entire patient time series of data. This made their approach nearly optimal, against which GLUCOSE’s performance was measured. In reality, clinicians also only have access to data up to the current time step, making GLUCOSE ‘s performance particularly notable in this context. This suggests that GLUCOSE can provide a valuable tool in the management of hyperglycemia in this critically ill patient population, offering a level of reliability and clinical applicability that is on par with traditional human-driven dosing strategies. The ability of GLUCOSE to maintain high performance across diverse clinical scenarios further supports its potential integration into clinical practice, where it could enhance patient outcomes by reducing variability in insulin dosing and minimizing the risks associated with both hyperglycemia and hypoglycemia.

Incorporation of distributional RL is another significant strength of this study. Even among patients with seemingly similar clinical profiles, there can be considerable variation in physiological responses. Distributional RL is particularly well-suited to address this challenge, as it quantifies the intrinsic uncertainty within a Markov Decision Process (MDP), which is characteristic of stochastic environments (18). By learning to approximate the distribution of potential outcomes, this approach strengthens the model by preparing it to handle the inherent uncertainties of real-world clinical settings. While GLUCOSE demonstrates significant potential, several limitations should be considered when interpreting these results. First, although our retrospective study strongly supports the use of GLUCOSE as a clinical decision support tool, these findings require validation through prospective studies and clinical trials involving large and diverse patient cohorts. Second, the GLUCOSE model has been trained, tested, and externally validated only for the first day following cardiac surgery. Though this is the most dynamic time-period for patients after cardiac surgery, expanding this work to evaluate insulin regimens over longer postoperative periods would further enhance GLUCOSE’s clinical utility. Third, our current algorithm does not incorporate dietary data. In practice, precise dietary data is seldom collected, and cardiac surgery patients often have restricted dietary intake during the first postoperative day, reducing the relevance of this variable. Prior work has also shown that RL algorithms can achieve successful blood glucose control without explicit dietary information (44). GLUCOSE learns underlying patterns associated with meals rather than relying on direct meal data, making its implementation more practical by reducing the need for collecting non-routine, precise dietary information. Finally, to ensure accurate training and validation of the model, we restricted ourselves to patients that had accurately documented doses of medications such as insulin, given that it was the action, and vasopressors/inotropes, which indicate the risk of disease severity and thus may portend a higher risk of hyperglycemia, in both development and external validation cohorts. With this, we did not need to exclude any patients in MIMIC-IV, but had to exclude 4,838 patients in eICU-CRD dataset. This missingness in eICU-CRD is a known issue with the dataset (29), but with patients from over 200 hospitals, it is a highly heterogenous dataset and thus remains impactful for external validation. To ensure that our model’s recommendations still generalize appropriately in the excluded subset of the external validation dataset we assessed the distribution of recommendations of the model in the subset with just ambigious insulin data, and in the subset with ambiguous data about both insulin and vasopressor/inotropes. We found that the distribution of recommendations was very similar in the 3 groups, with no clinically meaningful differences in actions, which suggests good generalizability of the model.

In summary, we have developed and externally validated GLUCOSE, a distributional RL based model to dynamically optimize glucose management in cardiac surgery patients. The comprehensive three-phase human evaluations support GLUCOSE’s clinical robustness and safety, demonstrating its effectiveness in real-world settings and its performance on par with or surpassing that of experienced clinicians. Future studies should be focused on randomized controlled trials to further evaluate the effectiveness and safety of GLUCOSE in diverse clinical settings.

## METHODS

### Study Design and Databases

For this retrospective study, we used the MIMIC-IV database (20) to develop the GLUCOSE algorithm (Development dataset). MIMIC-IV is a single-center database constructed from deidentified ICU admissions at the Beth Israel Deaconess Medical Center from 2008-2019. We externally validated the derived policy using the heterogeneous eICU Collaborative Research Database (eICU-CRD) (21) (External Validation Dataset). eICU-CRD is constructed from over 200,000 de-identified admissions to 208 United States hospitals between 2014 and 2015.

### Study Population

We included all adult patients (age ≥ 18 years) who were admitted to ICU after cardiac surgery. We used ICD-9-PCS and ICD-10-PCS codes to identify patients who underwent cardiac surgery in MIMIC-IV database (Supplementary Table 3). The eICU-CRD database does not include ICD-9-PCS or ICD-10-PCS procedure codes. As per prior literature (21), we have identified patients admitted to the ICU after cardiac surgery using the “admissiondx” table that provides the primary diagnosis for ICU admissions (Supplementary Table 4). We excluded patients who died within first 24 hours of ICU admission, had ambiguous medication administration information such that it did not allow us to calculate the exact dose of medication administered, or did not have available glucose levels within first three hours of documented ICU admission time after surgery. As our focus was to develop a policy to personalize the administration of regular insulin, we excluded patients who received other short acting insulins (aspart, lispro, NPH, insulin 70/30) (Supplementary Fig. 3).

### Feature Extraction and Preprocessing

We extracted information about patient demographics (age, sex, race), comorbidities (history of diabetes, hypertension, end stage renal disease, chronic obstructive pulmonary disease, asthma, prior myocardial infarction, congestive heart failure, Elixhauser comorbidity score), laboratory values (complete blood count, comprehensive metabolic panel, coagulation studies, and blood gases), vital signs (systolic blood pressure, diastolic blood pressure, mean arterial pressure, heart rate, respiratory rate, temperature, and oxygen saturation), vasopressor and inotrope doses, mechanical ventilation status, and SOFA scores. We extracted the data as multidimensional discrete time series in 1-h time intervals, with features summed or averaged as clinically appropriate. We excluded features with over 30% missingness. In line with standard approach to handling missingness in these data, we used forward fill imputation for all features with *k*-nearest neighbor (*k*-NN) imputation (*k*=5) to impute any remaining missing data (23, 46). Only the first 24 hours of data for each patient was utilized. All features were checked for outliers using a frequency histogram and descriptive statistics. Errors were corrected as appropriate, such as conversion of temperature to Fahrenheit to Celsius. The full feature list can be found in the Supplement Table 1. All features across all datasets were normalized into range [0, 1] based on the training set to improve training stability.

Our outcome was appropriate glucose control, defined as an hourly glucose level between 70-180mg/dL (10, 47), in the first day after cardiac surgery. Consequently, we began recording timesteps from the availability of the first glucose level measurement after admission to ICU.

### Computational Modeling

We used conservative Q learning (CQL), a state-of-the-art offline RL algorithm that allows model to suggest clinical actions while regularizing the learned policy to mitigate overestimation in low-coverage or out-of-distribution state-action pairs (48). To further enhance the model’s understanding of uncertainty and risk, we integrated CQL with distributional RL, an approach that captures the entire distribution of returns rather than just the expected return. All models use a multi-layer perceptron (MLP) network with 3 512-dimension hidden layers. This integration is crucial for making safe and effective decisions especially with clinical actions that have a narrow therapeutic index, such as insulin dosing. To achieve this, we incorporated Implicit Quantile Networks (IQN) into CQL, leveraging the strengths of distributional RL to better model the variability in patient responses, thereby improving the robustness of GLUCOSE (19). Unlike other distributional methods that require explicitly defining the number of quantiles, IQN implicitly models the entire return distribution, providing a more comprehensive representation of potential outcomes while improving upon its non distributional counterparts (17–19). To the best of our knowledge, this is the first application of integration of CQL with distributional Q functions in healthcare.

Finally, we implemented a batch training sampling strategy for offline RL, which avoids overregularization by low-return actions, allowing the learned policy to reflect more high-return trajectories (49).

### State Space

RL typically considers problems as MDPs. An MDP can be represented as a tuple of (*s_t_, a, r, s_t+1_)* for each time step *t*. Here, *s*_t_ represents a vector observation of features at that hour index *t*, and *s_t+1_*represents the the state at the next hour index after taking action *a.* The reward, *r*, is given for taking action *a* at state *s_t_*.

We used the features derived from demographics, comorbidities, laboratory values, vital signs, medications, mechanical ventilation, and SOFA scores binned into hourly time-steps to develop the state space. Based on previous literature, we incorporated the prior four hours of glucose values, when available, into the RL model (50). To provide additional context, we included information on glucose level changes during this period and calculated the ratio of glucose change to insulin dose for each hour, with the minimum insulin dose set at 0.1 for this calculation.

### Action Space

In our offline RL model, actions are defined as the amount of regular insulin administered each hour, utilizing a continuous action space. For ease of interpretability, we have rounded the recommended insulin doses to the nearest integer. This practice aligns the model’s recommendations with practical clinical standards and facilitates the clinical implementation of suggested doses. Additionally, we capped the insulin doses recommended by GLUCOSE at a maximum of 10 units per hour. This threshold is more than two standard deviations above the mean administered dose of 2.2 units in MIMIC-IV. We selected this cap due to both the low coverage of higher doses in the training data and to mitigate potential safety risks associated with recommending excessively high doses.

### Reward

We designed our reward to maximize optimal glycemic control while strongly discouraging behavior that would result in both hypoglycemia (glucose < 70 mg/dL) and hyperglycemia (glucose > 180 mg/dL). We provided a maximum reward of +0.2 within the 140-180 mg/dL range and penalties become increasingly negative, down to to -1, outside that range. We chose a reward with relatively low magnitude to improve training stability, and a relatively negative reward to disincentive any out of range values (51).

The reward is outlined in Equation 1:

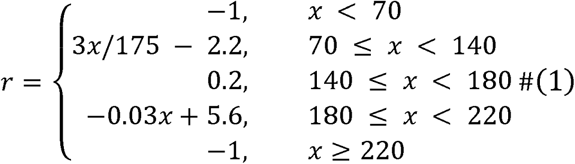

To ensure the safety of insulin dosing recommendations, considering insulin’s narrow therapeutic window, we implemented an exponentially increasing penalty that serves to discourage large overcorrections and promotes more cautious dosing adjustments (52):

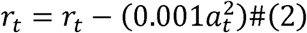

### GLUCOSE Model Training and External Validation

To train the policies, we first split the development dataset into 85% training and 15% internal testing sets. Since RL is particularly subject to high stochastic training variation (22), we sought to mitigate sampling and stochastic biases by training multiple models on subsampled 80% splits of the training set. Each training run and sampling split utilized a unique seed to ensure different training sets and distinct sampling order while maintaining reproducibility. We continued training models until substantial and significant improvements over the clinician policies were observed in the 15% internal testing set. The final model (GLUCOSE) was selected as the one that had the highest lower bound of the 95% CI for estimated performance returns in the internal testing set (23). We then evaluate GLUCOSE on the external validation set (Fig. 1).

Training was conducted in batches of 256, with actor and critic learning rates of 1e^-4^ and 3e^-4^, respectively. The discount factor γ was set at 0.67, corresponding to a 3 hour effective horizon (calculated as 1/(1-γ)). Discount factors are problem specific, and the choice of a lower discount factor is critical in the context of this problem. Higher discount factors, such as those exceeding 0.95, extend the effective horizon beyond the episode length, resulting in future rewards being weighted nearly as heavily as immediate rewards. Glucose levels can change rapidly, which could lead to suboptimal policy development as the model may either overly prioritize distant future rewards unaffected by the current state or become insensitive to immediate low reward states. Using a lower discount factor aligns the temporal focus of the model to ensure it remains responsive to rapidly changing glucose levels. A dropout rate of *p*=0.1 was applied during training to improve policy generalization. All models were implemented in Python 3.8.2 using d3rlpy (53).

### Off-Policy Evaluation

We used fitted-Q-evaluation (FQE) for offline policy estimation (OPE) to estimate the performance of the learned policies (26, 27). FQE is effective in handling large policy deviations from observed behaviors as well as stochasticity, and it has shown consistency and calibration in various healthcare-specific benchmarks (54, 55). Bootstrapping was applied across all episodes in the datasets to generate 95% confidence intervals by sampling with replacement 10,000 times. We then estimated the performance using FQE on both internal testing set and external validation dataset.

We further explored policy performance by analyzing the time in range (TIR) (70-180 mg/dL) in relation to the difference of GLUCOSE’s dosing recommendations and clinician-administered doses (Fig. 2). We calculated the cumulative differences as the model’s predicted insulin doses minus the observed insulin doses over the first 24 hours of ICU stay:

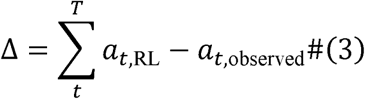

Cumulative differences are positive when the RL model recommends more insulin than what was administered, while negative differences indicate that the model predicted a lower insulin dose compared to what was observed.

### Estimation of Feature Importance

The interpretability of machine learning models is crucial in clinical care, where the rationale behind a model’s predictions must be clear to ensure patient safety and informed decision-making. SHAP, a method grounded in cooperative game theory, assesses the contribution of each feature to a model’s prediction by analyzing all possible combinations of feature values (28). In this study, we employ Permutation SHAP to estimate these contributions, as it provides a model-agnostic framework for elucidating model outputs.

### Human Evaluations

We further assessed the clinical validity of GLUCOSE in three separate phases of human evaluations. In the first phase, we compared the hourly insulin dosing recommended by GLUCOSE to that administered to patients in both internal testing and external validation datasets. Two senior endocrinologists (RS, AG), each with over ten years of clinical experience, provided their own hourly insulin dosing recommendations for 10 randomly selected patients in each cohort. Using the average hourly insulin doses recommended by endocrinologists as reference, we compared the insulin doses recommended by GLUCOSE to those administered to these patients.

In the second phase, we had two senior cardiac intensivists (>5 years’ experience) (PM, VN), two junior cardiac intensivists (<5 years’ experience) (DP, JG), and two cardiac intensive care unit nurse practitioners (AC, DG) provide their recommendations for hourly insulin doses in the first day after cardiac surgery for the same patients. We then compared the GLUCOSE recommended doses to those recommended by these clinicians, again using the average endocrinologist recommendations as references.

In the third phase, a panel of two senior intensivists (AS, GK) conducted a blinded evaluation of GLUCOSE against other clinician recommended insulin dosing schemes. Senior intensivists were selected for this phase because, in practice, these frontline clinicians are frequently responsible for making rapid decisions regarding glucose control in critically ill patients. They evaluated each dosing scheme for each patient using the following 5-point Likert scale questions:

*1.* Q1 (Safety) - How much risk for hypoglycemia does this regimen put the patient at? 1. Very high risk 2. High risk 3. Medium risk 4. Low risk 5. Minimal risk
*2.* Q2 (Effectiveness) - How effective is this regimen in bringing the glucose level within an acceptable range? 1. Not effective at all 2. Slightly effective 3. Moderately effective 4. Very effective 5. Extremely effective
*3.* Q3 (Acceptability) - How acceptable would this regimen be to you in clinical settings? 1. Strongly unacceptable 2. Unacceptable 3. Neutral 4. Acceptable 5. Strongly acceptable

### Evaluation of Model Recommendations in Excluded Patient Subsets

We also evaluated the GLUCOSE ‘s recommendations using the part of external validation dataset that was excluded from the primary analysis due to presence of ambiguous insulin administration data, which prevented us from making direct comparisons or calculating OPE. Patients for which exact doses of insulin, vasopressors, or inotropes could not be resolved were separated and underwent the same exclusion criteria and preprocessing used for the primary external validation cohort. Any ambiguous data was zero-filled.

### Statistical Analysis

We performed comparisons of categorical features using Chi-squared test and continuous features using t-test and Kruskal-Wallis test, as appropriate. All significance levels were set at α=0.05. To compare insulin doses administered by clinicians and GLUCOSE before hypo- and hyperglycemic episodes, we used Mann-Whitney U tests given the skewed distributions. To evaluate the accuracy of the insulin dosing schemes, we calculated the mean absolute error (MAE) between the insulin doses recommended by various dosing schemes to those provided by endocrinologists. MAEs were determined by subtracting the endocrinologists’ recommendations from the doses suggested by clinicians and the GLUCOSE system for each hourly dose administered or recommended for the 20 retrospectively reviewed patients. We then performed a t-test to identify any significant differences in MAE between the observed clinicians’ dosing and the endocrinologist’s suggested dosing, and MAE between GLUCOSE’s suggested dosing and the endocrinologist’s suggested dosing. To assess differences in the average insulin doses across subsets of excluded patients, we used ANOVA tests for group-wise assessment and two sided t-tests for pairwise assessment. All statistical tests were conducted using Python 3.8.2 using SciPy (56).

## Data Availability

All datasets are available online as below -
Study uses data from MIMIC-IV (https://physionet.org/content/mimiciv/3.1/) and eICU-CRD (https://eicu-crd.mit.edu) datasets

## ACKNOWLEDGEMENTS

This study was supported by NIH/NIDDK grant 5K08DK131286 (AS). This work was supported in part through the computational and data resources and staff expertise provided by Scientific Computing and Data at the Icahn School of Medicine at Mount Sinai and supported by the Clinical and Translational Science Awards (CTSA) grant UL1TR004419 from the National Center for Advancing Translational Sciences. Research reported in this publication was also supported by the Office of Research Infrastructure of the National Institutes of Health under award number S10OD026880 and S10OD030463. The content is solely the responsibility of the authors and does not necessarily represent the official views of the National Institutes of Health.

