## Supplementary for "GLUCOSE: A Distributional Reinforcement Learning Model for Optimal Glucose Control After Cardiac Surgery"

**Supplementary Table 1:**

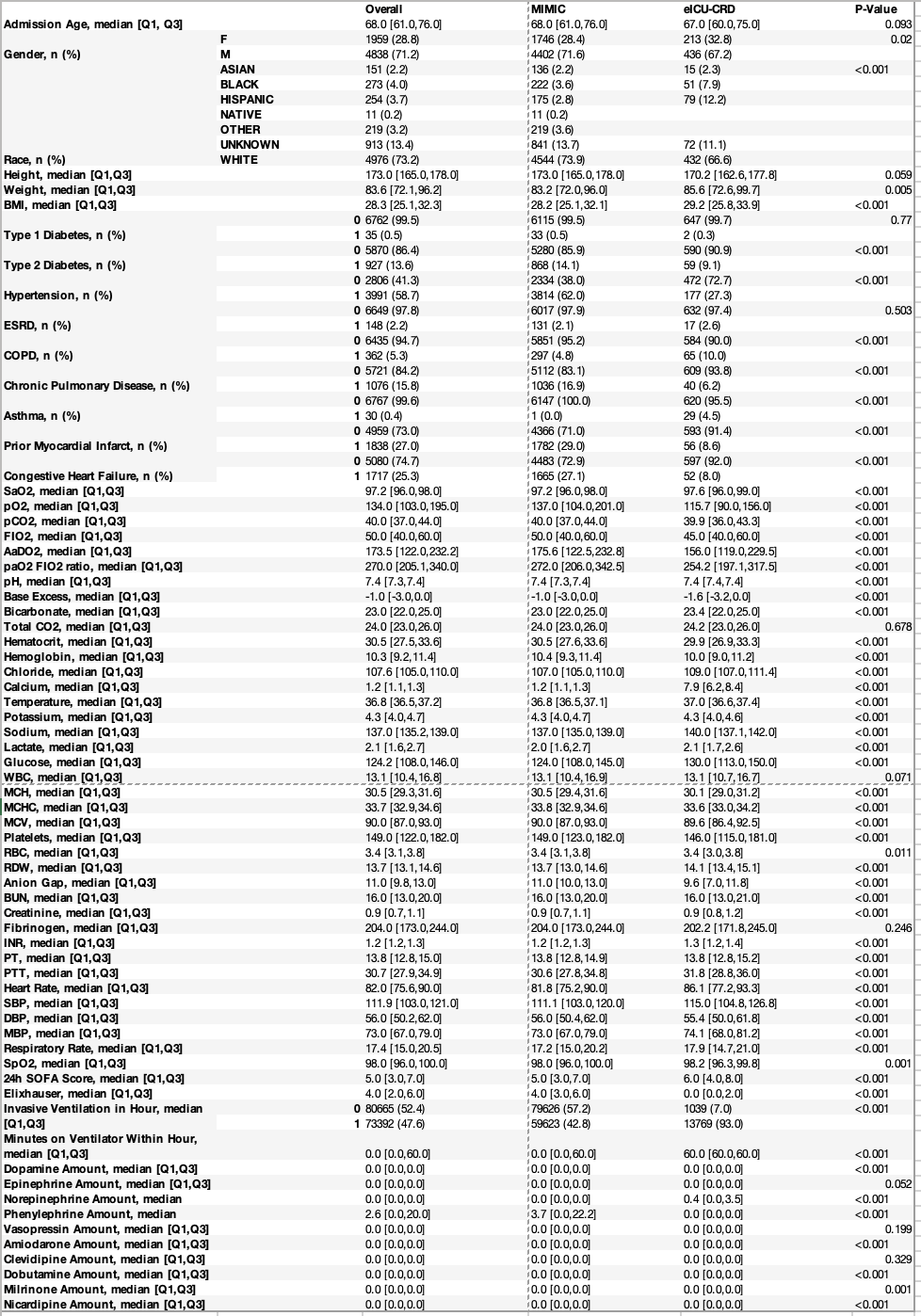

**Supplementary Table 2:**

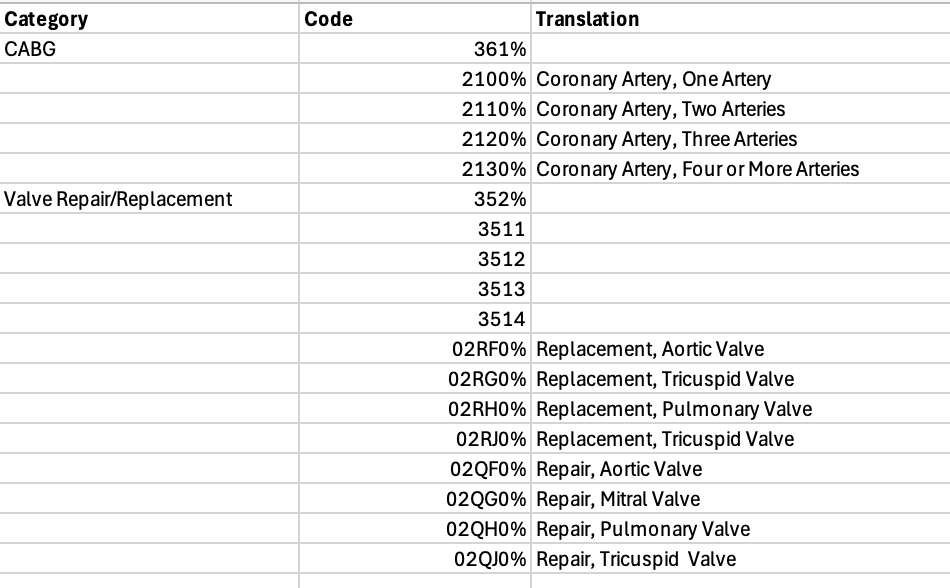

**Supplementary Table 3:**

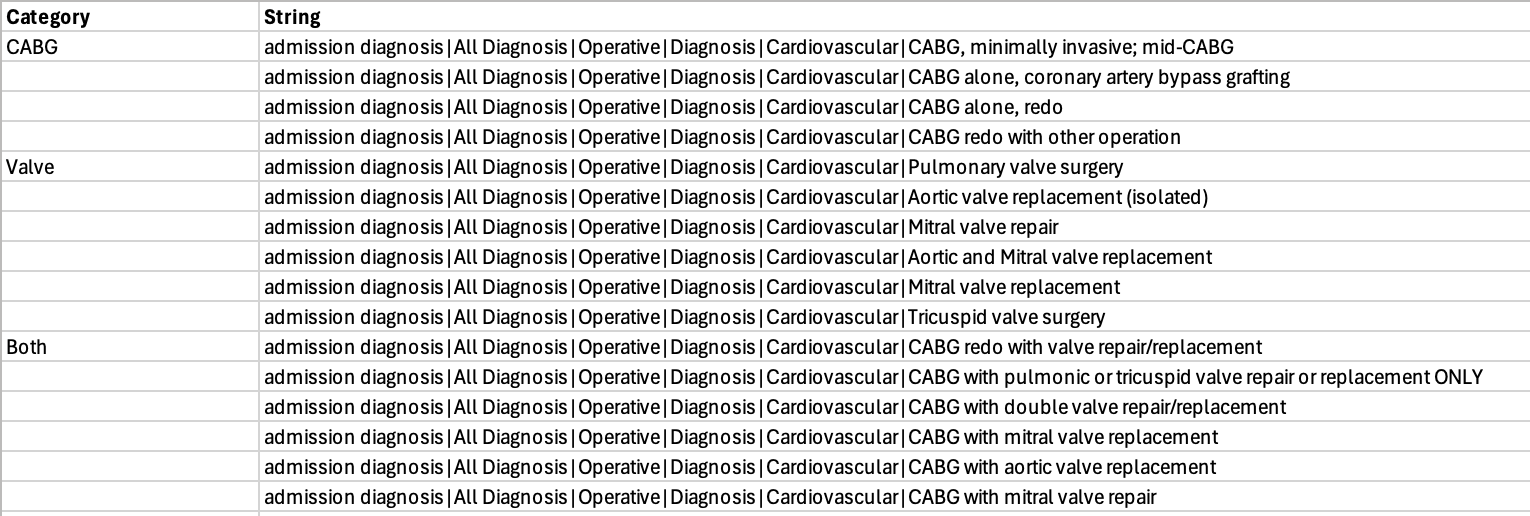

**Supplementary Figure 1:** Feature Importance by SHaP

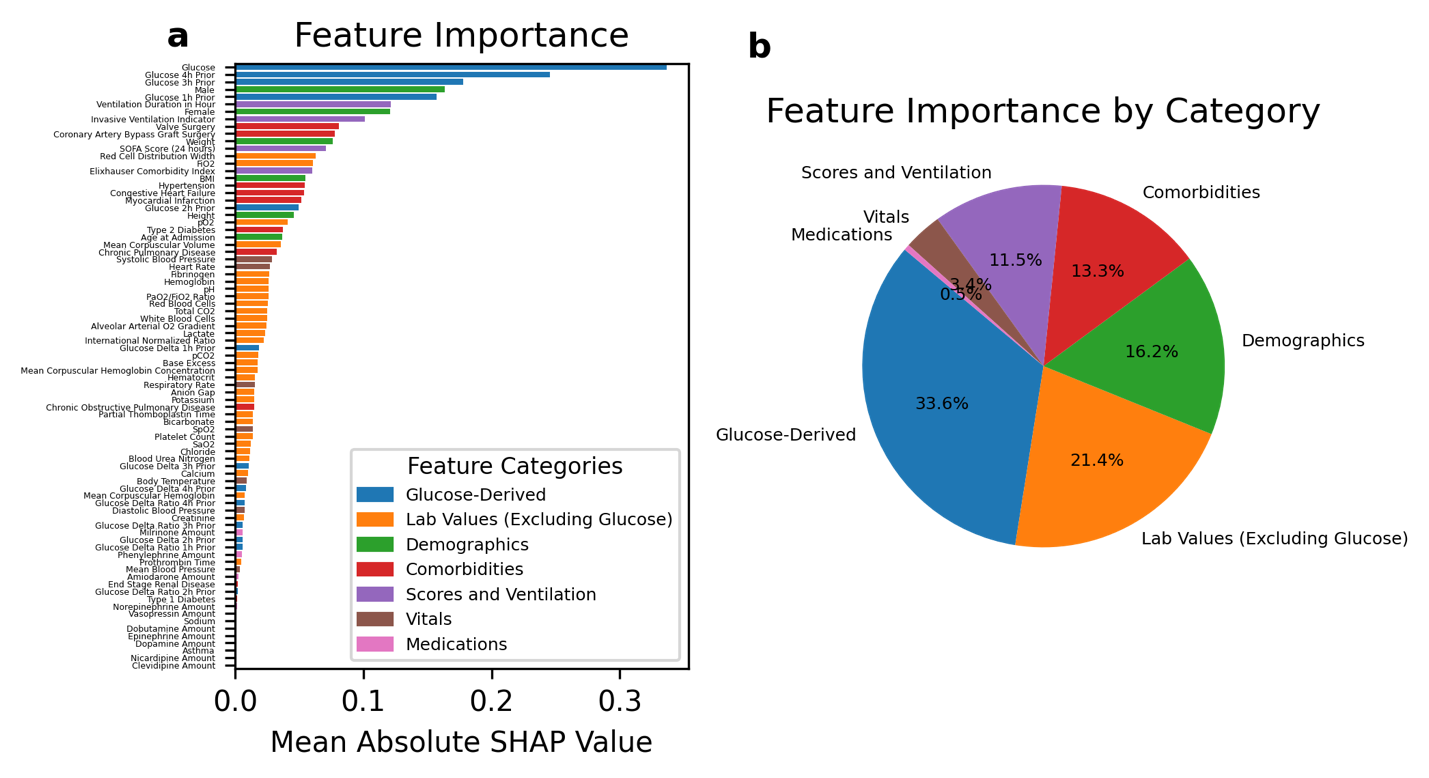

**Supplementary Figure 2:** Study Inclusion and Exclusion Criteria

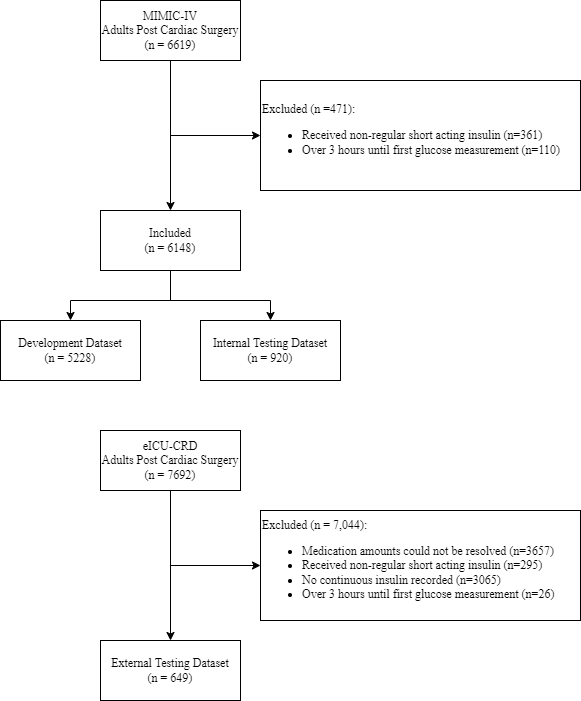
